# An Individualized Tractography Pipeline for the Nucleus Basalis of Meynert Lateral Tract

**DOI:** 10.1101/2023.08.31.23294922

**Authors:** Rachel A. Crockett, Kevin B. Wilkins, Michael M. Zeineh, Jennifer A. McNab, Jaimie M. Henderson, Vivek P. Buch, Helen M. Brontë-Stewart

## Abstract

**Background:** At the center of the cortical cholinergic network, the nucleus basalis of Meynert (NBM) is crucial for the cognitive domains most vulnerable in PD. Preclinical evidence has demonstrated the positive impact of NBM deep brain stimulation (DBS) on cognition but early human trials have had mixed results. It is possible that DBS of the lateral NBM efferent white matter fiber bundle may be more effective at improving cognitive-motor function. However, precise tractography modelling is required to identify the optimal target for neurosurgical planning. Individualized tractography approaches have been shown to be highly effective for accurately identifying DBS targets but have yet to be developed for the NBM.

**Methods:** Using structural and diffusion weighted imaging, we developed a tractography pipeline for precise individualized identification of the lateral NBM target tract. Using dice similarity coefficients, the reliability of the tractography outputs was assessed across three cohorts to investigate: 1) whether this manual pipeline is more reliable than an existing automated pipeline currently used in the literature; 2) the inter- and intra-rater reliability of our pipeline in research scans of patients with PD; and 3) the reliability and practicality of this pipeline in clinical scans of DBS patients.

**Results:** The individualized manual pipeline was found to be significantly more reliable than the existing automated pipeline for both the segmentation of the NBM region itself (p<0.001) and the reconstruction of the target lateral tract (p=0.002). There was also no significant difference between the reliability of two different raters in the PD cohort (p=0.25), which showed high inter- (mean Dice coefficient >0.6) and intra-rater (mean Dice coefficient >0.7) reliability across runs. Finally, the pipeline was shown to be highly reliable within the clinical scans (mean Dice coefficient = 0.77). However, accurate reconstruction was only evident in 7/10 tracts.

**Conclusion:** We have developed a reliable tractography pipeline for the identification and analysis of the NBM lateral tract in research and clinical grade imaging of healthy young adult and PD patient scans.

## Introduction

Disability and death due to Parkinson’s disease (PD) is increasing faster than any other neurological disorder (1). While the cardinal motor symptoms of PD, bradykinesia, rigidity, and tremor (2) can be improved by dopaminergic interventions, treatments for cognitive impairment remain scarce and ineffective. The nucleus basalis of Meynert (NBM) is at the center of the cortical cholinergic network and is crucial for the cognitive domains most vulnerable in PD (3). Preclinical evidence has demonstrated the positive impact of NBM deep brain stimulation (DBS) on cognition (4,5) but early evidence in humans has had mixed results (6–10). It is possible that the lateral NBM efferent white matter fiber bundle may be a novel and more effective DBS target for treating cognitive-motor syndrome. However, precise tractography modelling is required to identify the optimal target for neurosurgical planning.

The NBM is involved in tasks requiring attention, visuospatial, and executive functions (3). It is known to degenerate early in PD, which was predictive of cognitive but not motor impairment up to five years later (11). While reduced integrity of the NBM region has been shown to predict cognitive impairment in patients with PD (12), reduced integrity of the NBM white matter fiber bundles was shown to have a stronger contribution to cognitive function than the NBM region itself (13–15). There are two main pathways emanating from the NBM: 1) a lateral pathway that travels through the external capsule and uncinate fasciculus to supply the frontal, insula, parietal, and temporal cortices; and 2) a medial pathway, passing through the cingulum to supply the parolfactory, cingulate, pericingulate and retrosplenial cortices (16). Our previous work has shown greater mean diffusivity and reduced fractional anisotropy of both the lateral and medial NBM tracts in patients with early stage PD up to one year prior to the development of mild cognitive impairment (17). However, only the lateral tract has been associated with cognitive outcomes in older adults with mild cognitive impairment and dementia (14).

To date, DBS of the NBM in humans with Alzheimer’s disease (8,9), dementia with Lewy bodies (7), and PD (6,10) has primarily targeted the grey matter nucleus itself with varying levels of symptom improvement. Evidence from the clinical depression literature indicates that targeting specific white matter fiber bundles may be more effective than exclusively focusing on grey matter nuclei targets. While targeting grey matter targets has been shown to be beneficial in approximately 60% of patients with depression (18), Riva-Posse and colleagues (19) demonstrated that using individualized tractography to guide neurosurgical planning of the target of interest resulted in 82% of patients responding to DBS with over 50% in remission after 12 months. Similarly, tractography guided DBS of the ventral intermediate nucleus of the thalamus was also found to be more successful at treating essential tremor, than targeting the nucleus directly (20). Thus, tractography guided approaches for DBS may have broad benefit.

Current methods for recreating the lateral NBM tract uses automated segmentation of regions of interest (13,14). Both of the most commonly used NBM masks from healthy adults were segmented by a probabilistic mask determined from magnetic resonance imaging and histology from between one to ten post-mortem brains (21,22). However, the reliability of using masks developed in post-mortem histology slices to in-vivo neuroimaging has been questioned (23). This is particularly challenging for small subcortical brain regions without very high scan resolution or stark contrast differences. In addition, these masks are developed from very small samples, which may not be account for inter-subject variability, especially in clinical populations with a high level of atrophy. Thus, there is a need for more reliable individualized approaches to identify the NBM region and tract reconstruction. George and colleagues (24) have developed a manual approach for segmenting the substantia innominata (SI), which contains the NBM, using structural neuroimaging. However, their approach was specific to volumetric analysis of the SI and did not separate the NBM from the surrounding anatomy. A manual tractography approach for reconstructing the lateral NBM tract has yet to be evaluated.

Therefore, the aim of this study is to expand on these methods by using multi-modal neuroimaging and an individualized manual approach to develop an improved tractography methodology for the reconstruction of the NBM lateral tract. We aim to assess the reliability of our approach in three ways. First, we compare our manual pipeline to that of the existing automated pipeline used in the literature (13) in a test-retest sample of high-quality research scans from healthy young adults. Second, given the manual nature of our proposed approach, we aim to confirm the reproducibility of its use across researchers by assessing the inter- and intra-rater reliability in research quality scans from a cohort of patients. Finally, to investigate the use of this pipeline in shorter clinical DTI scans, we will determine the test-retest reliability in a cohort of DBS patients from our database. The development of this pipeline will be beneficial both for more reliable investigation of the cortical cholinergic network as well as to guide the neurosurgical planning for DBS procedures that aim to target this region.

## Methods

### Participants

#### Cohort 1

Thirty-seven participants from the test-retest cohort from the Human Connectome Project (HCP) (25) were included in the first stage of our analyses. Participants were included in the HCP study if they were: 1) aged 22-35 years old; 2) had no significant history of psychiatric disorder, substance abuse, neurological or cardiovascular disease; 3) had no evidence of cognitive impairment as indicated by a score ≥29 on the Mini Mental State Examination (MMSE); and 4) were able to provide informed consent. They were excluded if they: 1) had a history of seizures or epilepsy; 2) had a genetic disorder such as cystic fibrosis; 3) had been taking prescription medications for migraines in the last 12 months; 4) had multiple sclerosis, cerebral palsy, brain tumor, stroke, sickle cell disease, or experienced a severe head injury; 5) were currently on chemotherapy or immunomodulatory agent or had a history of this treatment that could affect the brain; 6) were currently being treated for diabetes; 7) were a premature birth; or 8) did not meet the safety criteria to undergo neuroimaging.

#### Cohort 2

Twenty-one participants with PD from the Parkinson’s Progression Markers Initiative (PPMI) database (www.ppmi-info.org/access-data-specimens/download-data, RRID:SCR_006431) were included in the second stage of this study. For up-to-date information on the PPMI study, visit www.ppmi-info.org. Participants were included in the PPMI study if they: 1) were ≥30 years old; 2) had PD for at least 2 years prior to screening; 3) were not currently on or expected to start PD medication for at least 6 months after baseline testing; 4) were not currently or planning to become pregnant during the study; and 5) were able to provide informed consent. They were excluded if they: 1) were currently taking PD medication; 2) had other atypical PD syndromes (e.g., metabolic disorders, encephalitis, or other neurodegenerative disease); 3) were clinically diagnosed with dementia; or 4) had any other medical or psychiatric condition which might preclude participation. Additional inclusion criteria for the current study required participants to have: 1) undergone magnetic resonance imaging (MRI) with both a structural T1-weighted (T1-w) and diffusion weighted (DWI) scan; and 2) have evidence of mild cognitive impairment at baseline indicated by a score of 23-25 on the Montreal Cognitive Assessment (MoCA), which is consistent with the criteria for a patient to be eligible for NBM DBS to treat cognitive impairment.

#### Cohort 3

Five patients (aged 64 – 74 years, 1 female) with PD from our ongoing research investigating DBS of the subthalamic nucleus were included in the final clinical cohort. Participants were included in the parent study if they: 1) were over 18 years of age; 2) had a diagnosis of idiopathic PD; 3) had documented improvement in motor symptoms with dopaminergic medication; and 4) were on stable doses of medication. They were excluded if they: 1) were over 80 years of age; 2) had significant cognitive impairment or dementia as determined by a standardized neuropsychological battery; 3) were clinical depressed as defined by the DSM-IV; 4) had very advanced stage PD as indicated by Hoehn and Yahr stage 5 while on medication; 5) did not meet the requirements for MRI and/or neurosurgery; 6) had a history of seizures, or who required electroconvulsive therapy or transcranial magnetic stimulation to treat a chronic condition; 7) were unable to comply with study follow-up visits; or 8) were unable to provide informed consent.

### Magnetic Resonance Imaging (MRI) Acquisition

#### Cohort 1

Data from the HCP were acquired using a Siemens 3T scanner with high-performance head-only gradients. The T1-w images consisted of 256 slices, with a 0.7mm^3^ voxel size, 224×224 field of view, 2400ms repetition time (TR), and 2.14ms echo time (TE). The diffusion tensor echo planar images (DTI) consisted of 111 slices, with 1.25mm^3^ voxel size, a 168 × 144 acquisition matrix, 5520ms TR, 89.5ms TE, 90 gradient directions with *b =* 1000, 2000, and 3000 s/mm^2^ and six *b*0 images.

#### Cohort 2

The MRI scans from the PPMI database were acquired using standardized procedures across sites on a Siemens 3T scanner. The T1-w images consisted of 192 slices, with a 1mm^3^ voxel size, a 256 × 256 acquisition matrix, 2300ms TR, and 2.98ms TE. The DTI consisted of ∼80 slices, with a of 2mm^3^ voxel size. They had a 128 × 128 acquisition matrix, ∼10000ms TR, ∼80ms TE, 64 gradient directions with *b* = 1000 s/mm^2^ and one *b*0 image, and an acquisition time of ∼ 95.5s.

#### Cohort 3

The clinical MRI scans were acquired on a Discovery MR750 3T scanner at the Stanford Neuroscience Health Center. The T1-w image consisted of 188 slices, with a 1mm^3^ voxel size, 256 × 256 acquisition matrix, ∼ 8.51ms TR and ∼ 3.2ms TE. The DTI consisted of one 31 slice scan encoded with 30 gradient directions in the posterior to anterior direction and another consisting of two slices with one gradient in the anterior to posterior direction. They had a 2mm^3^ voxel size, 128 x128 acquisition matrix, ∼ 8000ms TR, 61ms TE, *b* = 1000 s/mm^2^ with one *b*0 image for each scan, and an acquisition time of 90.6s.

### Tractography Pipeline

Preprocessing of the MRI data was completed using FMRIB Software Library (FSL) (26). Prior to completing the clinical tractography pipeline, topup (27) and eddy (28) correction were used to correct for distortion and eddy currents respectively. The data in cohort 1 had already undergone these preprocessing steps (29). A diffusion tensor model was fit using FSL DTIFIT, producing colorized fractional anisotropy (FA) images. Removal of non-brain structures was completed using HD-BET (30) on the T1-w, DTI and the *b*0 slice. All images were then aligned to the anterior commissure-posterior commissure (ACPC) line using the rigid linear conversion from T1-w to the MNI ACPC line with six degrees of freedom as the reference transformation (31). The gradient direction matrices (bvecs) were also rotated to account for this transformation.

For both pipelines, the brainstem was extracted using FSL’s First automated segmentation tool (32), and the hemisphere mask was hand drawn in FSLeyes by identifying the midline of the brain in the coronal plane on the subject scans.

### Manual Segmentation of Regions of Interest

The optic tract and anterior commissure were identified using both the T1-w and FA maps and manually segmented using FSLeyes. The anterior end of the anterior commissure mask was identified by the slice at which there was a clear band across the coronal plane and was segmented through its full posterior extent.

The external capsule was identified in the axial plane of the DWI (see Figure 1). The most inferior slice began where there is a clear outline of the internal capsule that is not overlapping with the anterior commissure. From this slice, the external capsule is drawn stopping slightly short of the anterior and posterior ends of the capsule. To avoid the tractography being guided by overlapping tracts in the more superior portions of the external capsule, the superior boundary was specified at approximately 9mm (7 slices for scans with a 1.25mm voxel size) above the inferior slice.

**Figure 1.**
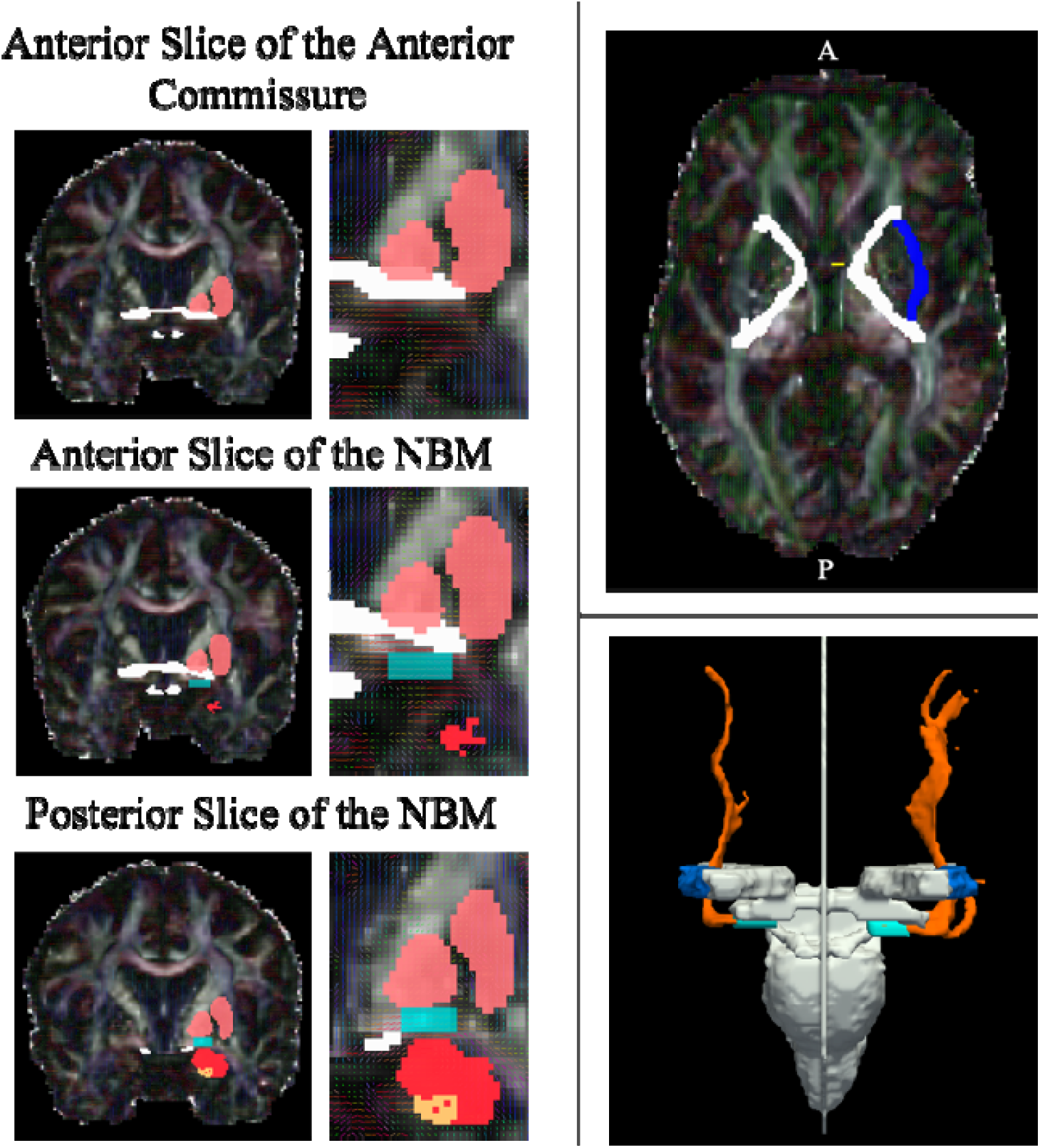
Left: Examples of the anterior and posterior slices of the NBM mask (cyan) using the pallidum/putamen (pink), anterior commissure and optic tract (white), amygdala (red), and hippocampus (orange) as guidance regions. Top Right: Example mask of the inferior slice of the internal capsule (white) not overlapping with the anterior commissure (yellow) and encapsulating the left external mask (navy). Bottom Right: Example tractography output (orange) with the seed NBM (cyan), external capsule (navy) and exclusion regions (white).

The internal capsule mask is then identified across the same axial slices ensuring that the anterior and posterior ends of the external capsules are sealed off by the internal capsule mask.

Finally, the NBM masks were identified in the coronal plane using a combination of the manually drawn anterior commissure and optic tract, and the automated extraction of the pallidum and hippocampus from the FSL First segmentation, overlaying the outputted primary eigenvector image onto the FA image. The NBM was identified by a strong lateral (red) directionality of the voxels below the pallidum and within the guidance anatomy (see Figure 1). The most anterior slice of the anterior commissure (the point at which there is a strong and clear band across the coronal slice) was used to identify the most anterior portion of the NBM. Starting at least one slice posterior to this marker, signified the first slice of the anterior NBM. In scans where identification of the anterior commissure was challenging, the point at which the inferior end of the fornix column meets the base of the internal capsule was used as an additional indicator of the anterior NBM boundary. The posterior slice was indicated at the point whereby the hippocampus begins to become evident. The medio-lateral border of the NBM was aligned with the medial and lateral edges of the pallidum. The anterior commissure and optic tract masks were used to avoid overlapping regions of the NBM.

### Automated Segmentation of Regions of Interest

The automated tractography pipeline was designed based on methods previously used for identifying the NBM tracts of interest in older adults with and without neurodegenerative disease (13,14).

All regions of interest were identified using predetermined atlases in MNI space, which were then transformed to subject space. The internal capsule, external capsule, and cingulum were extracted from the Johns Hopkins University white matter atlas (33), the anterior commissure from FSL’s XTRACT atlas (34), and the NBMs from the Ch4 basal forebrain regions identified by stereotaxic probabilistic maps from 10 high quality post-mortem brains (21). Consistent with the appropriate threshold for realistic NBM extraction (23), the probabilistic NBM masks were threshold at 50%.

### Probabilistic Tractography

After preprocessing the data with bedpostX, probabilistic tractography was completed using the standard 5000 samples and 0.2 curvature threshold settings in probtrackX with the NBM as the seed region, and the ipsilateral external capsule as the waypoint region of inclusion. The anterior commissure, internal capsule, brain stem, and hemisphere were specified as regions of avoidance. The extracted tracts were threshold at 90% and binarized.

To compare the similarity of the shape of the binarized tracts, Dice similarity coefficients were calculated using fsl_dice. Dice similarity coefficients are a metric of how identical two binary images are (35). A Dice coefficient equal to one is indicative of complete overlap while a score of zero indicates no overlap. For comparisons whereby the tracts were extracted from two separate scans, the DTIs from both runs were co-registered and the midpoint of the transformation used to create a halfway point as a common space across the two scans. The tracts and NBM masks from each run were then transformed into this space to allow for structural comparisons using Dice coefficients.

### Data Analyses

For cohort 1, the spatial overlap of the tracts and NBM masks were compared using Dice coefficients between data processed separately from each run for the HCP dataset. A paired-samples t-test was used to investigate differences in Dice coefficient between the manual and automated pipeline on the HCP data. For cohort 2, Dice coefficients were also used to investigate the reliability of the manual pipeline on PD research scans. Paired-samples t-tests were used to compare the within-rater Dice coefficients across researchers, and the between-rater Dice coefficients across runs. Finally, for cohort 3, Dice coefficients were used to investigate the reliability of the manual approach for recreating the tracts of interest in clinical scans of people with PD. An alpha of 0.05 was used to indicate statistical significance.

## Results

### Cohort 1: Manual vs Automated in Healthy Young Adult Research Scans

Two extreme outliers (Mean ± 3SD) were identified in the Dice coefficient of the automated NBM masks. However, removal of these datapoints did not alter the significance of the results and thus were kept in the final analyses. The individualized manual pipeline had significantly higher Dice coefficient than the existing automated pipeline across the HCP test-retest cohort for both the NBM mask itself (t=4.24, p<0.001) and the lateral NBM tracts (t=3.17, p=0.002). This indicates higher test-retest reliability of the manual pipeline (see Figure 2).

**Figure 2.**
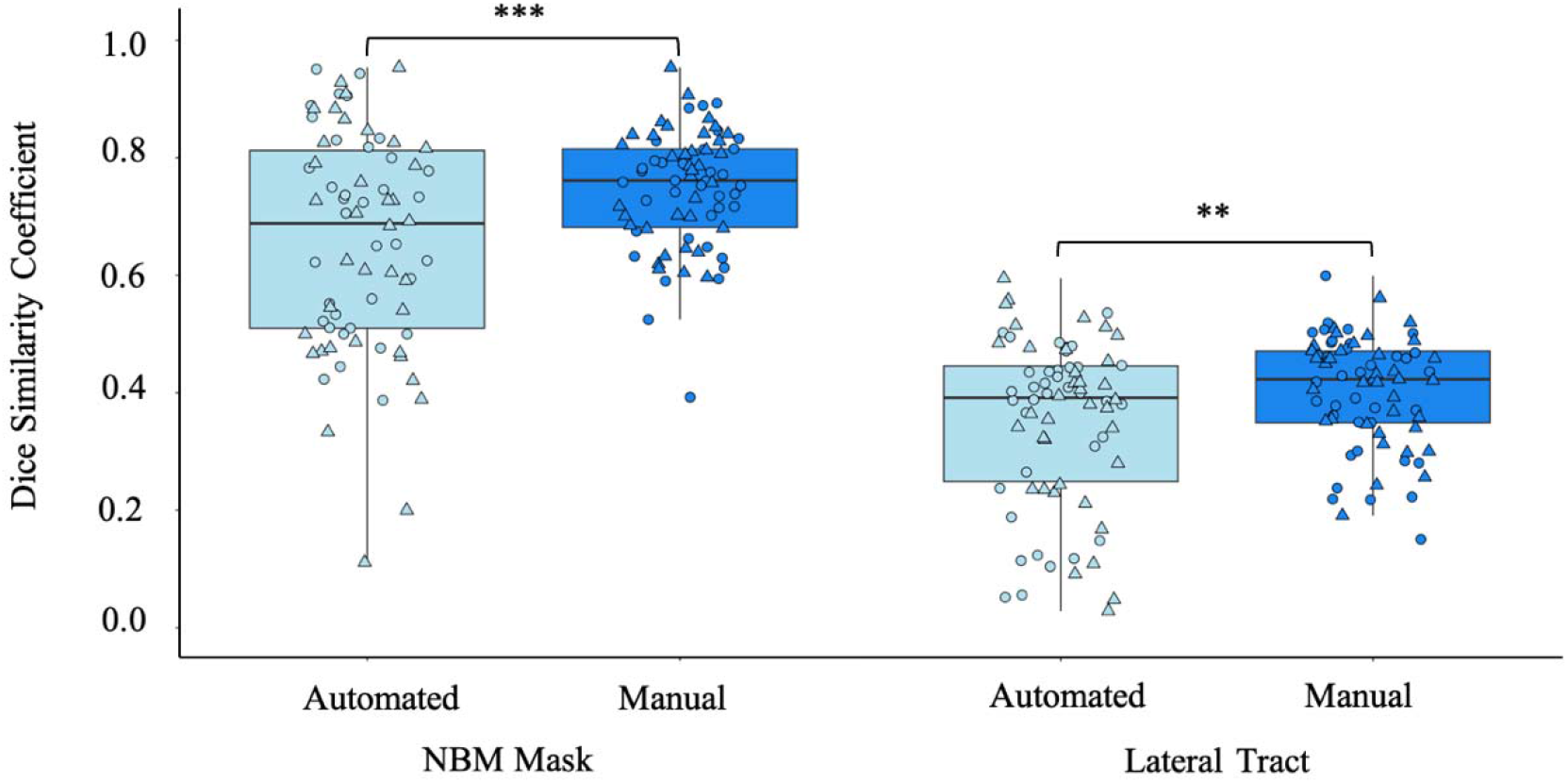
Comparison of the test-retest reliability of the automated and manual pipeline tractography outputs from healthy young adult high quality research scans. Circle: Left hemisphere; Triangle: Right hemisphere. **p<0.01, ***p<0.001

### Cohort 2: Inter- and Intra-Rater Reliability of the Manual Pipeline in Patients with PD

When running the pipeline on research quality scans in PD patients who may be eligible for NBM DBS, we found high Dice coefficients for both raters between runs (mean >0.70) indicating high intra-rater reliability. Notably, there was no significant difference (t=1.17, p=0.25) in the intra-rater Dice coefficient between raters suggesting both raters were equally reliable. In addition, there was no significant difference in the inter-rater reliability for each run (t=-0.31, p=0.75). The mean between-rater Dice coefficient was 0.65 for run 1 and 0.66 for run 2 indicating high inter-rater reliability of the pipeline (see Figure 3).

**Figure 3.**
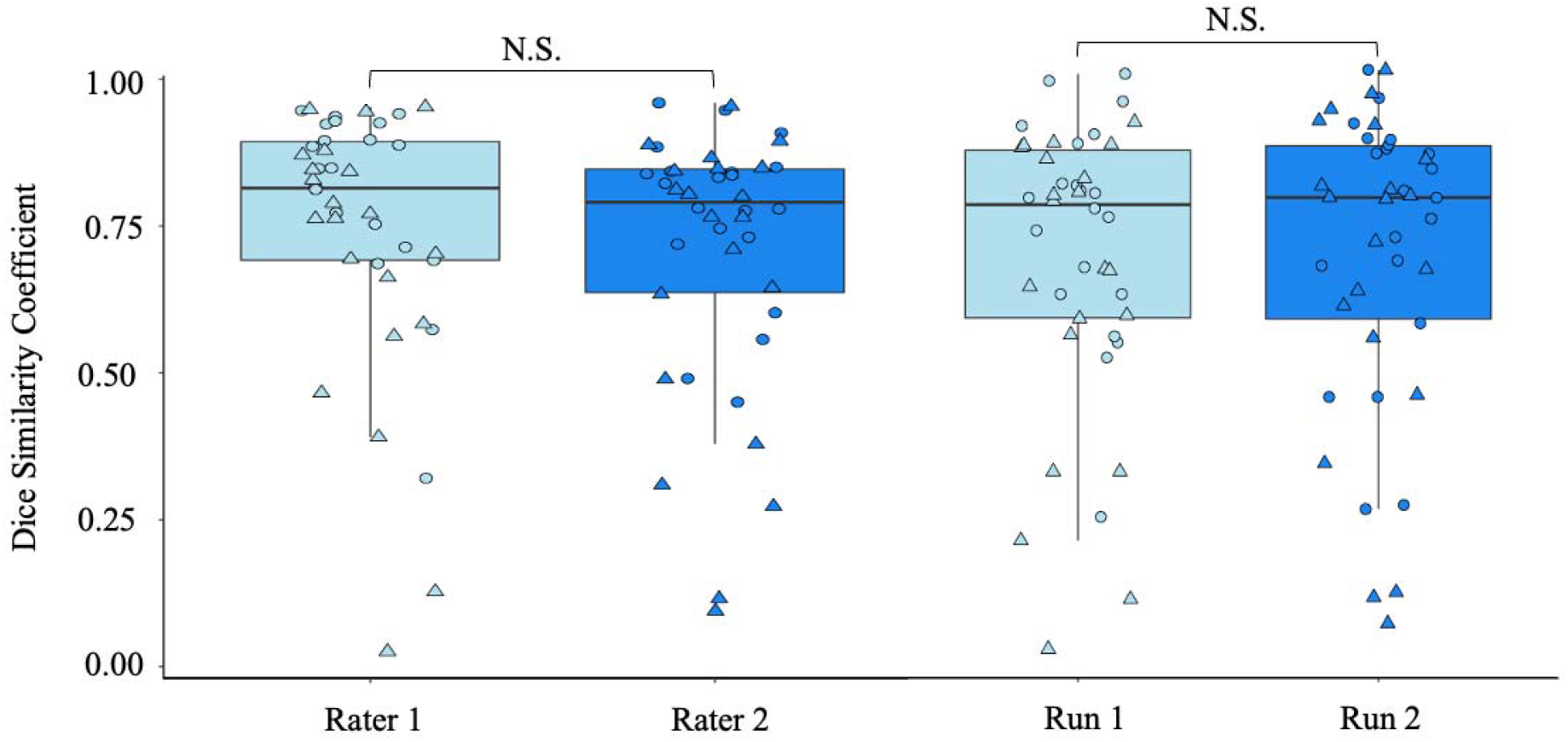
Left: comparison of the test-retest reliability for two different raters; and Right: comparison of the inter-rater reliability across each run using data from the Parkinson’s Progression Markers Initiative. Circle: Left hemisphere; Triangle: Right hemisphere; N.S. Not significant.

### Cohort 3: Reliability of Manual Pipeline in Clinical Scans

The mean (SD) Dice coefficient for the clinical scans was 0.77 (0.1), which is indicative of very high reliability across runs (see Figure 4). Visual inspection of the data demonstrated a 70% success rate for both runs in the reconstruction of the tract of interest. The target tracts were not successfully recreated across either run for both hemispheres in subject 2 and for the right side in subject 1.

**Figure 4.**
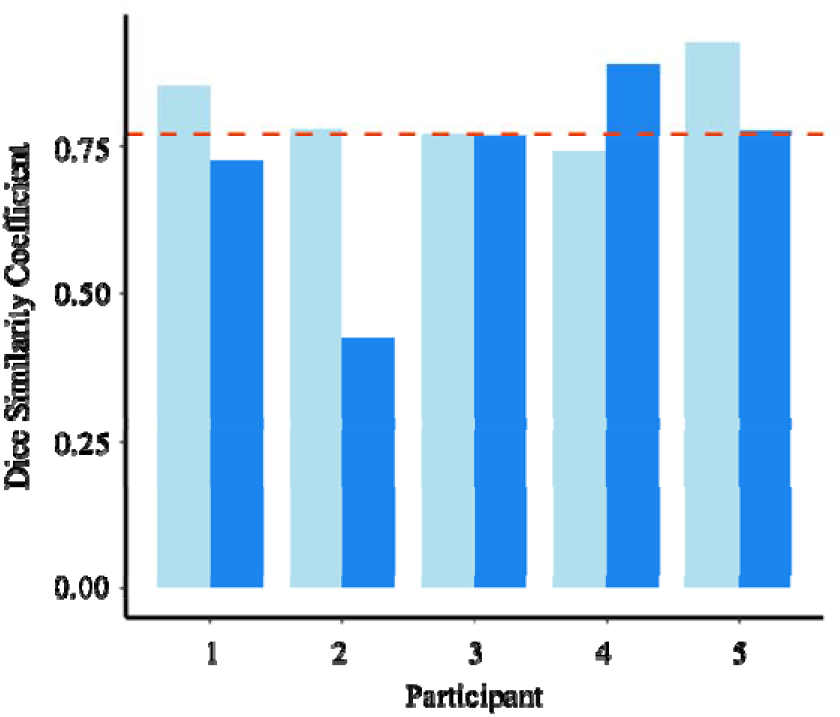
Dice similarity coefficients for the test-retest reliability of the manual tractography approach in clinical scans. Cyan: Left hemisphere; Blue: Right hemisphere. The dashed line indicates the mean Dice coefficient (0.77) across the full sample.

## Discussion

We successfully developed a novel individualized tractography pipeline for the reconstruction of the NBM lateral tracts. The pipeline was found to have greater test-retest reliability than existing automated methods in high quality MRI scans of healthy young adults. We also found high inter- and intra-rater reliability across research and clinical grade MRI scans in patients with PD who are eligible for DBS. However, greater consideration maybe needed when using this approach in clinical neuroimaging of patients with high levels of atrophy and lower quality scanning parameters.

Previous methods to identify and recreate the NBM tracts have used automated methods based on small samples of post-mortem brains leading to inconsistent results (23). More recently, Doss and colleagues (36), used a deep learning methods to improve the accuracy of automated segmentation approaches. However, this approach still relied on the initial manual identification of the NBM in a healthy cohort, which was then translated to a clinical population of temporal lobe epilepsy patients. We expanded on the manual segmentation work by George and colleagues (24) by narrowing the anatomical focus to that of the NBM exclusively and using both structural and diffusion weighted imaging for enhanced visualization of the region of interest. It is notable that manually identifying the NBM was more effective when combined with the manual identification of the anterior commissure rather than using existing automated masks which struggled to identify this region as effectively. The anterior commissure is also used as a region of exclusion within the probabilistic tractography pipeline. Therefore, the manual segmentation of this region has the additional benefit of providing greater guidance for the NBM tract reconstruction. Overall, we identified that our manual individualized pipeline recreated more consistent NBM masks and subsequently the NBM lateral tracts than using automated mask segmentations, indicating improved test-retest reliability. It is notable that the overall inter-scan Dice coefficient for the HCP cohort was lower than when assessing the reliability of the pipeline in the same scan for the PPMI and DBS patient cohorts. Consequently, the reliability of both approaches is slightly reduced when inter-scanner/repeat scanning is a factor.

While the manual individualized approach may lead to more precise tractography outcomes, it can also be vulnerable to greater inter-rater variability. Thus, we sought to evaluate the reliability of this pipeline across two trained researchers. We identified high inter- and intra-rater reliability between researchers and across tractography runs respectively. Importantly, there was no significant difference between the dice coefficients comparing each rater across two runs of tractography. This indicates that both raters were equally reliable at recreating the tracts of interest. In addition, this analysis was shown to be reliable within a cohort of PD patients who would be eligible for NBM DBS (i.e., had mild cognitive impairment). Therefore, indicating the benefit of this approach for both healthy young adults and those with neurodegenerative disease.

Finally, we aimed to investigate the use of this tractography approach for clinical use. We identified high test-retest reliability in the clinical scans. However, there were three tracts which were not reproduceable for either run. This is likely a result of lower quality imaging parameters. For instance, compared to the 1.25mm^3^ voxel size, 90 gradient directions and three *b*0 values of the high-quality scans from the HCP, clinical MRI generally only use a 2mm^3^ voxel size, 30 gradient directions, and only one *b*0 value. This reduces the sensitivity of the scan to detect accurate fiber orientations. The smaller voxel size is also particularly relevant for spatial resolution given the very small size of the NBM (∼ 3 × 13.5 × 17mm) (37,38). Thus, the larger slice thickness in clinical scans may lead to either over or under estimation of the NBM region itself.

The need to develop improved research tools to examine the integrity of these major white matter tracts is of vital importance. A growing body of evidence suggests changes in white matter may be more sensitive at identifying differences in clinical populations than grey matter. Taylor and colleagues (39) showed that changes in white but not grey matter integrity was associated with changes in symptom severity in de-novo PD patients over 12 months. This supports the finding that deficits to the lateral NBM white matter fiber bundle, but not the grey matter nuclei itself, was predictive of cognitive decline in patients with Alzheimer’s disease or dementia with Lewy bodies as well (14). We have also highlighted the reduced integrity of the NBM white matter tracts up to one year prior to the development of cognitive impairment in PD patients (17). In addition to neurodegenerative disease, reduced integrity of the white matter was identified in patients with Euthymic bipolar I disorder compared to controls (40). However, no differences were identified between groups in the grey matter. These findings suggest a greater need for neuroimaging analysis tools that can unlock new white matter targets of investigation. The ability to expand the focus of future research will provide vital insight into the underlying neurobiology of a multitude of neurological diseases, which is crucial for early detection and treatment.

This study is not without limitations. Firstly, the lower Dice coefficient values across both pipelines in the HCP data suggests an impact of inter-scanner noise on the consistency of tractography outputs. We did not have repeat scans at the same timepoint for the PPMI and clinical data, which reduced our ability to investigate the test-retest reliability across different scanning procedures in the PD population. We also chose to only assess the reliability of these pipelines using the FSL probabilistic method of tractography. While this is considered the most robust method (41,42), we did not assess how these approaches may compare across other tractography algorithms, such as deterministic tractography, that may also be used in research and/or clinical settings. Lastly, at present, there is no ground truth for the identification of the lateral NBM tract. Therefore, the interpretation of these tract outputs is limited to visual comparison with histological maps of target tract. However, the nature of these tracts from these histological studies are well documented (16,37).

## Conclusion

Our novel individualized tractography pipeline was shown to be highly reliable at reconstructing the lateral NBM tracts in both healthy young adults and patients with PD. This pipeline can be used for improved reliability of research investigating the cortical cholinergic network in aging and neurodegenerative populations and can inform the neurosurgical planning of DBS aimed at targeting this network.

## Data Availability

Data from the Human Connectome Project can be requested from: www.humanconnectome.org/study/hcp-young-adult/data-releases
Data from the Parkinson's Progression Marker's Initiative can be requested from: www.ppmi-info.org/access-data-specimens/download-data

https://www.ppmi-info.org/access-data-specimens/download-data

## Author Contributions

RAC, KBW, and HBS were involved in study conception. RAC, KBW, MZ, and JM contributed to the methodological planning. RAC and KBW completed the data processing and analyses. All authors were involved in interpretation of the findings. RAC wrote the first draft of the manuscript and all authors critically reviewed and approved the manuscript.

## Funding

Data were provided in part by the Human Connectome Project, WU-Minn Consortium (Principal Investigators: David Van Essen and Kamil Ugurbil; 1U54MH091657) funded by the 16 National Institute of Health (NIH) Institutes and Centers that support the NIH Blueprint for Neuroscience Research; and by the McDonnell Center for Systems Neuroscience at Washington University.

PPMI – a public-private partnership – is funded by the Michael J. Fox Foundation for Parkinson’s Research and funding partners, including, Abbvie, AcureX, Allergan, Amathus, Therapeutics, Asap, Avid, Biogen, Bial Biotech, Biolegend, BlueRock Therapeutics, Bristol-Myers, Calico, Celgene, Cerevel, Coave Therapeutics, Dacapo BrainScience, Jenali Therapeutics, 4D Pharma plc, GE Healthcare, Edmond J. Safra Philanthropic Foundation, Genentech, GlaxoSmithKline, Golub Capital, Gain Therapeutics, Handl Therapeutics, Insitro, Janssen Neuroscience, Lilly, Lundbeck, Merch, MSD Veso Scale Discovery, Neuroscine Biosciences, Pfizer, Piramal Healthcare, Prevail Therapeutics, Roche, Sanofi Genzyme, Servier, Takeda, Teva, UCB, Vanqua Bio, Verily, Voyager Therapeutics, and Yumanity Therapeutics.

This work was also supported in part by Michael J Fox Foundation (9605) and NIH: National Institute of Neurological Disorders and Stroke (NINDS) grants (R21 NS096398) to HBS. The authors are funded by two NIH: NINDS research grants (RAC, KBW, JMH, and HMBS: UG3NS128150; HMBS: UH3NS107709).

## Conflict of Interest

JMH is a consultant for Neuralink, serves on the Medical Advisory Board of Enspire DBS, and is a shareholder in Maplight Therapeutics.

## Notes

### Author Declarations

Data used in the preparation of this article were obtained from the Human Connectome Project (https://www.humanconnectome.org/study/hcp-young-adult/data-releases) and Parkinson's Progression Markers Initiative (PPMI) database (www.ppmi-info.org/access-data-specimens/download-data). For up-to-date information on the study, visit www.ppmi-info.org. For the cohort 3 clinical data referenced in this project, the IRB of Stanford University gave ethical approval for this work.

